# Data-driven subtypes of Alzheimer’s Disease: A multicohort study

**DOI:** 10.64898/2026.09.09.26362683

**Authors:** Amelie Metz, Cassandra Polizzi, Roqaie Moqadam, Katherine Chadwick, Cécilia Tremblay, Zhaojin Chen, Yashar Zeighami, Sylvia Villeneuve, the Alzheimer’s Disease Neuroimaging Initiative, the ARTFL-LEFFTDS Longitudinal Frontotemporal Lobar Degeneration (ALLFTD) Research Consortium, the Consortium for the early identification of Alzheimer’s Disease–Quebec (CIMA-Q), The PREVENT-AD Research Group, Mahsa Dadar

## Abstract

**INTRODUCTION:** Individuals with Alzheimer’s disease (AD) exhibit substantial heterogeneity in the severity of vascular changes and neurodegeneration, making the identification of biologically meaningful subtypes across the AD spectrum critical for improving prognosis and tailoring interventions. We aimed to determine whether data-driven subtyping of individuals at risk for or diagnosed with AD, based on magnetic resonance imaging (MRI), can identify distinct trajectories of disease progression and clinical outcomes.

**METHODS:** We analyzed baseline biomarkers and longitudinal clinical and cognitive assessments from the PREVENT-AD, CIMA-Q, ALLFTD, and ADNI cohorts, including 1396 participants. Regional atrophy and White Matter Hyperintensity (WMH) burden were derived from T1-weighted MRI. The SuStaIn algorithm was applied to baseline data to infer individuals’ subtypes and stages based on MRI biomarker profiles. The resulting subtypes were compared in terms of baseline molecular biomarker levels and vascular risk factors, longitudinal cognitive decline trajectories, and clinical and neuropathological outcomes.

**RESULTS:** Two distinct subtypes were identified across all cohorts: one characterized by early parahippocampal and cortical atrophy, and a second by initially elevated WMH volumes. Both subtypes showed abnormal tau and amyloid levels in- and ex-vivo, and subtype two exhibited increased vascular risk factors. Longitudinally, the atrophy-first subtype exhibited steeper cognitive decline than the WMH-first subtype. Accordingly, the atrophy-first subtype carried a higher risk of conversion to AD than the WMH-first group.

**DISCUSSION:** These findings, replicated in four diverse and independent datasets, support imaging-based subtyping as a scalable and clinically translatable approach for capturing heterogeneity in AD pathophysiology and associated cognitive trajectories.

## 1. Introduction

Alzheimer’s disease (AD) is the leading cause of dementia, affecting over 55 million individuals globally, and its prevalence is expected to double in the two decades as populations age.^1^ Pathological hallmarks include amyloid-β (Aβ) plaques, neurofibrillary tau tangles, and extensive neurodegeneration.^2^ Cerebrovascular damage represents a common co-pathology,^3^ frequently manifesting as white matter hyperintensities (WMH) on magnetic resonance imaging (MRI).^4–6^ Recent literature suggests disease-specific patterns of WMHs, with anterior WMHs reflecting vascular dementia, and posterior WMHs observed more frequently in AD.^7–9^

Despite a well-established biomarker cascade described in the AT(N) framework,^3^ beginning with Aβ accumulation (A), followed by tau propagation (T) and ultimately neurodegeneration (N), and frequently accompanied by vascular damage (V), studies have revealed substantial heterogeneity in the regional distribution and burden of proteinopathy and atrophy, even among individuals with comparable clinical severity.^10–14^ In a meta-analysis, Ferreira et al. (2020) synthesized evidence from 64 studies employing biological subtyping, either based on post-mortem histology or in-vivo imaging, and identified four subtypes. The most prevalent subtype, typical AD, was defined by tau-related pathology and atrophy in the hippocampus and association cortices. The limbic-predominant subtype was characterised by medial temporal lobe vulnerability and enriched for female sex, whereas the hippocampal-sparing subtype, with mostly neocortical involvement, presented at younger ages and with steeper cognitive decline. Between-subtype differences were further observed across cognition and fluid biomarker profiles, underscoring that biological subtypes reflect distinct pathophysiological entities with clinical differences.

A limitation of the conventional subtyping methods applied in these studies, such as hierarchical clustering, is the tendency to group disease subtypes based on overall disease burden rather than qualitatively distinct disease patterns. To overcome this, more recent work has adopted trajectory-based subtyping frameworks, like the Subtype and Stage Inference (SuStaIn) algorithm^15^, which simultaneously estimates subtype membership and disease stage. Implementations of this model revealed distinct subtypes of atrophy progression or tau deposition, respectively, corresponding to divergent clinical profiles and conversion rates.^15,16^ Recently, Mastenbroek et al (2025) integrated global measures of cortical atrophy, WMH burden, CSF Aβ and alpha-synuclein levels, tau PET signal, and ventricular volume in SuStaIn and identified five subtypes, with higher rates of cognitive decline in the pure AD and vascular groups, followed by the mixed AD and vascular subtype. Although the multimodal approach is a strength of this study, the authors relied exclusively on global measures, prohibiting the analysis of regional anatomical patterns that have proven informative in the aforementioned literature.

A further limitation of previous studies is their reliance on single-cohort samples which are inherently prone to bias, as participant eligibility criteria, recruitment strategies, imaging and testing protocols, and demographic composition vary substantially. This might obscure true patterns and constrain the generalizability of results. To address this, we harmonised data from four open-access, multicentre cohorts spanning North America, the Pre-symptomatic Evaluation of Novel or Experimental Treatments for Alzheimer’s Disease (PREVENT-AD)^17^, the Consortium for the early identification of Alzheimer’s disease-Quebec (CIMA-Q)^18^, the Alzheimer’s Disease Neuroimaging Initiative (ADNI)^19^, and ARTFL-LEFFTDS Longitudinal Frontotemporal Lobar Degeneration (ALLFTD), incorporating multimodal data for 1396 participants. Each cohort offers distinct and complementary characteristics. Notably, PREVENT-AD enrolled individuals with a first-degree family history of AD^17^, while CIMA-Q enriched their cohort with individuals with subjective cognitive decline (SCD)^18^. The ADNI on the other hand included an above-average proportion of APOE ε4 positive individuals^20^. Together, these datasets span the entire clinical spectrum from cognitively unimpaired to dementia, enabling cross-stage comparisons.

We applied a trajectory-based subtyping algorithm, SuStaIn, to baseline MRI data to identify clusters of individuals with similar biomarker profiles, while simultaneously estimating the temporal ordering of pathological changes. To capture co-pathological contributions, we included both regional atrophy and WMH burden. The resulting subtypes were then characterized in terms of their Aβ/tau burden, vascular risk factors, and longitudinal cognitive trajectories, with the aim of further uncovering the biological underpinnings of clinical heterogeneity in the AD spectrum.

## 2. Methods

### 2.1. Datasets

We included a total of 1396 participants from four open-access cohorts based on the availability of neuroimaging data. For the subtyping analysis, all participants with baseline T1-weighted MRI were included. For longitudinal follow-up analyses, we included participants who additionally had baseline and at least one follow-up assessment available (see sample numbers in Supplementary Figure 1). All studies were approved by the institutional review boards of participating institutions, and written informed consent was obtained from all participants or their authorized representatives.

#### 2.1.1. Alzheimer’s Disease Neuroimaging Initiative (ADNI)

The Alzheimer’s Disease Neuroimaging Initiative (ADNI, <u>adni.loni.usc.edu</u>)^19,20^ was launched in 2003 with the primary objective to determine whether serial MRI, positron emission tomography (PET), other biological markers, and clinical and neuropsychological assessments can be combined to measure the progression of MCI and early AD. Detailed inclusion and exclusion criteria are available at www.adni-info.org. Notably, participants were required to have a Hachinski Ischemic Score ≤ 4 to minimize inclusion of individuals with significant vascular contributions to cognitive impairment. We included 573 participants from ADNI-1, ADNI-2, ADNI-GO, and ADNI-3 cohorts, with available MRI at baseline.

#### 2.1.2. ARTFL-LEFFTDS Longitudinal Frontotemporal Lobar Degeneration (ALLFTD)

ARTFL-LEFFTDS Longitudinal Frontotemporal Lobar Degeneration (ALLFTD) is a multi-site study consisting of data collected from 23 North American institutions, which is a combination of two previously separate longitudinal neuroimaging studies, ARTFL and LEFFTDS (https://www.allftd.org/, https://memory.ucsf.edu/research-trials/research/allftd). It aims to longitudinally follow FTLD mutation carriers to improve understanding of FTLD disease progression based on both biological markers and clinical manifestation. Participants were primarily enrolled based on probable familial FTLD due to family history, but related sporadic disorders were also included. Data collected include MRI scans, fluid biomarkers, genetic testing, and clinical and cognitive variables. From the broader ALLFTD sample, we included 244 participants classified as AD, MCI, or cognitively healthy controls, restricting inclusion to those aged 55 years and above to ensure age-comparability with participants drawn from the other datasets.

#### 2.1.3. Consortium for the early identification of Alzheimer’s disease–Quebec (CIMA-Q)

We included 246 participants from the Consortium for the early identification of Alzheimer’s disease–Quebec (CIMA-Q).^18^ The main objective of CIMA-Q is the longitudinal characterization of an observational cohort of more than 422 elderly, cognitively healthy individuals with subjective cognitive disorders, suffering from mild cognitive disorders, or suffering from dementia due to probable AD. CIMA-Q collects clinical, cognitive, biological, radiological, and pathological data from these participants.

#### 2.1.4. Presymptomatic Evaluation of Experimental or Novel Treatments for Alzheimer’s Disease (PREVENT-AD)

We analyzed data from 333 participants in the Presymptomatic EValuation of Experimental or Novel Treatments for Alzheimer’s Disease (PREVENT-AD)^17^ cohort, data release 8.1 (https://www.centrestopad.com/), a longitudinal observational study of individuals with a first-degree family history of sporadic AD. PREVENT-AD was launched in 2011 and currently provides up to 12 annual visits consisting of multimodal MRI, PET, genetic, neurosensory, clinical, plasma/CSF, and cognitive testing data collected until 2024 on 348 participants.

### 2.2. MRI processing

T1-weighted structural MRI images were processed using the Pipeline for Evaluating Longitudinal Images of Cerebral Anatomy (PELICAN)^21^, an open-access pipeline built on the MNI MINC-Toolkit v2 and ANTs.^22–24^ Preprocessing included denoising,^25^ intensity inhomogeneity correction,^26^ intensity normalization, linear registration to the MNI-ICBM152 2009c template, followed by brain extraction with the BEaST algorithm^27^ and nonlinear registration using ANTs^24^. The BISON tool then segmented processed images into grey matter, white matter, and CSF probability maps.^28^ Nonlinear registrations and BISON maps were then used to generate voxel-based morphometry (VBM) maps quantifying regional grey matter volumes, as previously described^29^. Values for each cerebral lobe (frontal, temporal, parietal, occipital) as well as subcortical and parahippocampal regions were extracted using the CerebrA atlas^30^ for every participant. White matter hyperintensities (WMH) and ventricular volume were quantified using BISON. Lobar WMH volumes were extracted for each participant using the Hammers atlas^31,32^ and log-transformed to achieve a normal distribution. Linear and nonlinear registrations and segmentations were quality controlled by authors AM, RM, and KC, as previously described.^33^

### 2.3. Molecular markers

#### 2.3.1. ADNI

Fluid biomarkers included CSF t-tau, p-tau, and Aβ42 and plasma p-tau181. Tau measures were log-transformed to improve normality. Global amyloid burden was assessed using Florbetapir F-18 PET SUVRs, with cerebellar grey matter as the reference region. Acquisition and processing methods can be found at www.adni-info.org.

#### 2.3.2. ALLFTD

Fluid biomarkers included CSF t-tau and p-tau, and plasma p-tau217 and p-tau231. Tau measures were log-transformed to improve normality. Acquisition and processing methods are available upon request from the ALLFTD research study group.

#### 2.3.3. CIMA-Q

CSF biomarkers included t-tau, Aβ42, and Aβ40. Plasma biomarkers indexed global tau pathology using p-tau231. P-tau231 measures were log-transformed to improve normality. Acquisition and processing methods are further described in Belleville et al. (2019)^18^.

#### 2.3.4. PREVENT-AD

CSF biomarkers included total tau (t-tau), phosphorylated tau (p-tau), and amyloid-β42 (Aβ42). Plasma biomarkers indexed global tau pathology using p-tau231 and amyloid burden using Aβ42 and Aβ40. Tau measures were log-transformed to improve normality. Flortaucipir PET standardized uptake value ratios (SUVRs), calculated using inferior cerebellar grey matter as the reference region, were available for a meta-region of interest (meta-ROI) to quantify tau burden. Global amyloid burden was assessed using NAV4694 PET SUVRs, with cerebellar grey matter as the reference region. Acquisition and processing methods are further described in Villeneuve et al. (2025)^17^.

### 2.4. Vascular risk factors

We included vascular risk factors available in at least two of the included datasets. ADNI provided data on Body Mass Index (BMI) and diastolic and systolic blood pressure. ALLFTD provided data on BMI only. CIMA-Q provided data on BMI, diastolic and systolic blood pressure, total, high-density lipoprotein (HDL), and low-density lipoprotein (LDL) cholesterol, smoking status (including years smoking and cigarettes smoked per day), and diabetes. PREVENT-AD provided data on BMI, diastolic and systolic blood pressure, total, HDL, and LDL cholesterol, smoking status (including years smoking and cigarettes smoked per day), and diabetes.

For CIMA-Q, we also included volumetric measures of perivascular spaces (PVS) and microbleeds derived from T1w images. To this end, we applied SHIVA, a validated open-source PVS and microbleed segmentation technique.^34,35^ PVS were automatically segmented using T1w MRIs. Microbleeds were segmented using T2* MRIs. PVS segmentations were visually assessed to ensure their quality (author ZC). Microbleed segmentations were visually assessed and manually corrected (author MD) in cases of segmentation errors. PVS and microbleed counts were derived based on the total number of PVS and microbleeds segmented. PVS and microbleed volumes were calculated in the standard stereotaxic space (in mm^3^). Values were log transformed to achieve normal distribution.

### 2.5. Neuropsychological testing

To assess cognition, we included tests available in at least two datasets. Disease severity across all datasets was assessed using the Clinical Dementia Rating total score and sum of boxes. Global cognition was measured using the Mini-Mental State Examination (MMSE) (ADNI, CIMA-Q) and the Montreal Cognitive Assessment (MoCA) (ADNI, ALLFTD, CIMA-Q). Memory was assessed using the Rey Auditory Verbal Learning Test (RAVLT), including measures of verbal learning and immediate recall (ADNI, CIMA-Q, PREVENT-AD). Attention was evaluated using the Digit Span test (ADNI, ALLFTD, PREVENT-AD). Language and confrontation naming were assessed using the Boston Naming Test (BNT) (ADNI, CIMA-Q) and category verbal fluency (ADNI, ALLFTD, CIMA-Q). Across all cohorts, processing speed was measured using the Trail Making Test (TMT) Part A (time and number of errors), while executive function was assessed using TMT Part B (time and number of errors). PREVENT-AD and CIMA-Q also administered the Color-Word Interference Test (Stroop task) from the Delis-Kaplan Executive Function System, which assesses executive function under Inhibition and Switching conditions.

### 2.6. Neuropathological outcomes

Neuropathological confirmation was available for a subset of participants who underwent post-mortem brain autopsy (n = 38 in ADNI, n = 3 in CIMA-Q), following each cohort’s standardized neuropathological assessment protocol. Amyloid burden was characterized using Thal phase and CERAD neuritic plaque density score, while tau pathology was staged using Braak neurofibrillary tangle stage, with these measures combined into a composite ADNC (Alzheimer’s disease neuropathologic change) level. Cerebrovascular pathology was assessed through ratings of arteriolosclerosis, atherosclerosis of the circle of Willis, cerebral amyloid angiopathy (CAA) severity, and white matter rarefaction. Co-pathologies relevant to mixed dementia presentations were also captured, including presence of Lewy body related alpha-synucleinopathy, TDP-43 proteinopathy, and neuronal loss in the substantia nigra. Given the limited number of available specimens, continuous statistical analyses of these neuropathological measures were underpowered to detect meaningful associations. We therefore opted to categorize continuous or ordinal neuropathological variables using median splits, yielding three discrete levels: low (values below the sample median), medium (values equal to the median), and high (values above the median).

### 2.7. Subtype and Staging analysis

The Subtype and Stage Inference algorithm (SuStaIn)^15,36^ combines disease progression modelling with unsupervised machine learning to identify data-driven subtypes while accounting for disease stage. Unlike traditional methods that separate mixed cohorts (cognitively unimpaired to MCI) by overall severity, SuStaIn treats progression as piecewise-linear trajectories of w-scores (covariate-adjusted z-scores) across biomarkers. Biomarker features were w-scores for eleven variables: lobar white matter hyperintensity volumes (frontal, temporal, parietal, occipital), and regional gray matter VBM values (frontal lobe, temporal lobe, parietal lobe, subcortical areas, parahippocampal region). SuStaIn requires biomarker normalization relative to a cognitively normal reference distribution to account for effects of healthy aging, sex, education, and total intracranial volume (TIV). Normalization was based on the control groups of cognitively unimpaired individuals included in CIMA-Q, ADNI, and ALLFTD. For the subtype analysis, all datasets were merged and run together in one analysis. MRI outcomes were considered comparable between datasets as all sites followed the ADNI T1-weighted MRI protocol.

Based on the distributions of all markers, we defined three severity thresholds per biomarker (w = 0.7, 1.4, 2.1). The optimal number of subtypes was determined via 10-fold cross-validation, which evaluated model fit (log-likelihood) and cross-validation information criterion (CVIC). SuStaIn uses ten-fold cross-validation for two distinct purposes: (i) to evaluate the optimal number of subtypes and (ii) to evaluate the consistency of the subtype progression patterns. This enabled us to choose the model with the highest out-of-sample likelihood, or equivalently, the lowest value of the CVIC, across all folds. SuStaIn was fit to the full cohort using 10,000 Markov Chain Monte Carlo (MCMC) iterations for uncertainty diagnostic estimation, assigning each participant to their maximum-likelihood subtype and stage. Stage 0 is considered Subtype 0, meaning all w-scores are within the normal range.

### 2.8. Group comparisons

Analyses were conducted in R version 4.3.2. Baseline demographic characteristics across the identified subtypes were compared using one-way ANOVA for continuous variables and χ² tests for categorical variables. For significant ANOVA effects, post hoc pairwise comparisons were conducted with FDR correction. Given that data collection for fluid biomarkers, cognition, and vascular risk factors differed between datasets in terms of chosen markers or tests, the following group comparisons were conducted separately for each dataset. Baseline CSF, plasma, and PET marker levels for amyloid-beta and tau, as well as vascular risk factors, were compared across the resulting subtypes using linear regression models, adjusting for age and sex. Values were z-scored, and outliers beyond 4 standard deviations were excluded.

Neuropathological outcomes were compared across subtypes using multinomial logistic regression models, adjusted for age at death and sex, with results reported as odds ratios (ORs) compared to low levels of the respective pathological finding. Similarly, rates of clinical conversion to MCI or AD were compared across subtypes using logistic regression models, adjusted for age and sex, with stable healthy outcomes.

Subtype-specific cognitive decline trajectories were tested using linear mixed-effects models.

The model was specified as follows:

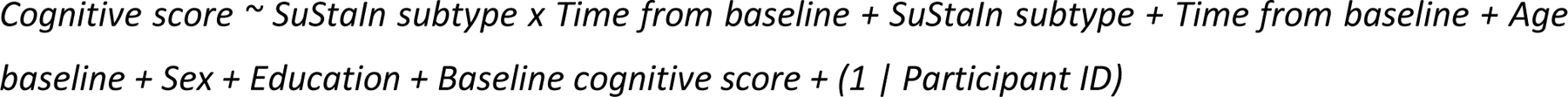

The SuStaIn subtype x time from baseline interaction tested whether the rate of cognitive decline differed across subtypes. Models controlled for age at baseline, sex, education, and baseline cognition, with categorical random intercepts per participant.

All values were z-scored, their directionality was adjusted so that lower values consistently correspond to worse performance, and outliers beyond 4 standard deviations were excluded. For longitudinal models, follow-up visits were cut at 3.5 years to ensure adequate sample sizes across subtypes. P-values were adjusted using false discovery rate (FDR) correction across tests and subtypes (p < 0.05)^37^. For the longitudinal assessments, only participants with at least 2 visits were included.

## 3. Results

### 3.1. Sample characteristics across datasets

The four cohorts included in this study differed in baseline age, sex, cerebrovascular disease burden, cognitive performance, and diagnostic distribution. While PREVENT-AD comprised only cognitively unimpaired individuals at recruitment, CIMA-Q included a higher proportion of participants with subjective cognitive decline, and ADNI had the greatest proportion of participants with MCI and AD (p < .001). Accordingly, MoCA scores were highest in PREVENT-AD and lowest in ADNI (p < .001). ADNI also included the highest proportion of APOE ε4 allele carriers, although APOE genotype was unavailable in ALLFTD. PREVENT-AD and ALLFTD were the youngest cohorts (p < .001). PREVENT-AD had a higher proportion of females than ALLFTD and ADNI (p < .01), whereas ADNI included more males than females. Total GM volume was highest in PREVENT-AD and CIMA-Q and lowest in ADNI (p < .001). WMH burden was greatest in ADNI and lowest in PREVENT-AD and ALLFTD (p < .001). Education levels did not differ significantly across cohorts (p = .052).

### 3.2. SuStaIn identifies two distinct subtypes

Cross-validation identified the two-subtype model as optimal (Supplementary Figure 3).

The two-subtype model resolved an atrophy-first and a WMH-first subtype in the merged sample (Figure 1A). The atrophy-first subtype (n = 448) was characterized by initial parietal and subcortical atrophy, followed by more widespread volume loss and increasing WMH burden at later stages. The regions reaching higher W-scores (W = 2.1), indicating more pronounced abnormality, earliest were the parahippocampal area and temporal lobe, consistent with the typical progression of neurodegeneration in AD. In contrast, the WMH-first subtype (n = 278) showed increased WMH burden first, beginning in the occipital lobe, followed by ventricular expansion and parahippocampal and temporal lobe atrophy. A total of 671 participants were classified as subtype 0, which SuStaIn considers healthy, i.e., their biomarkers were within the normal range.

**Figure 1.**
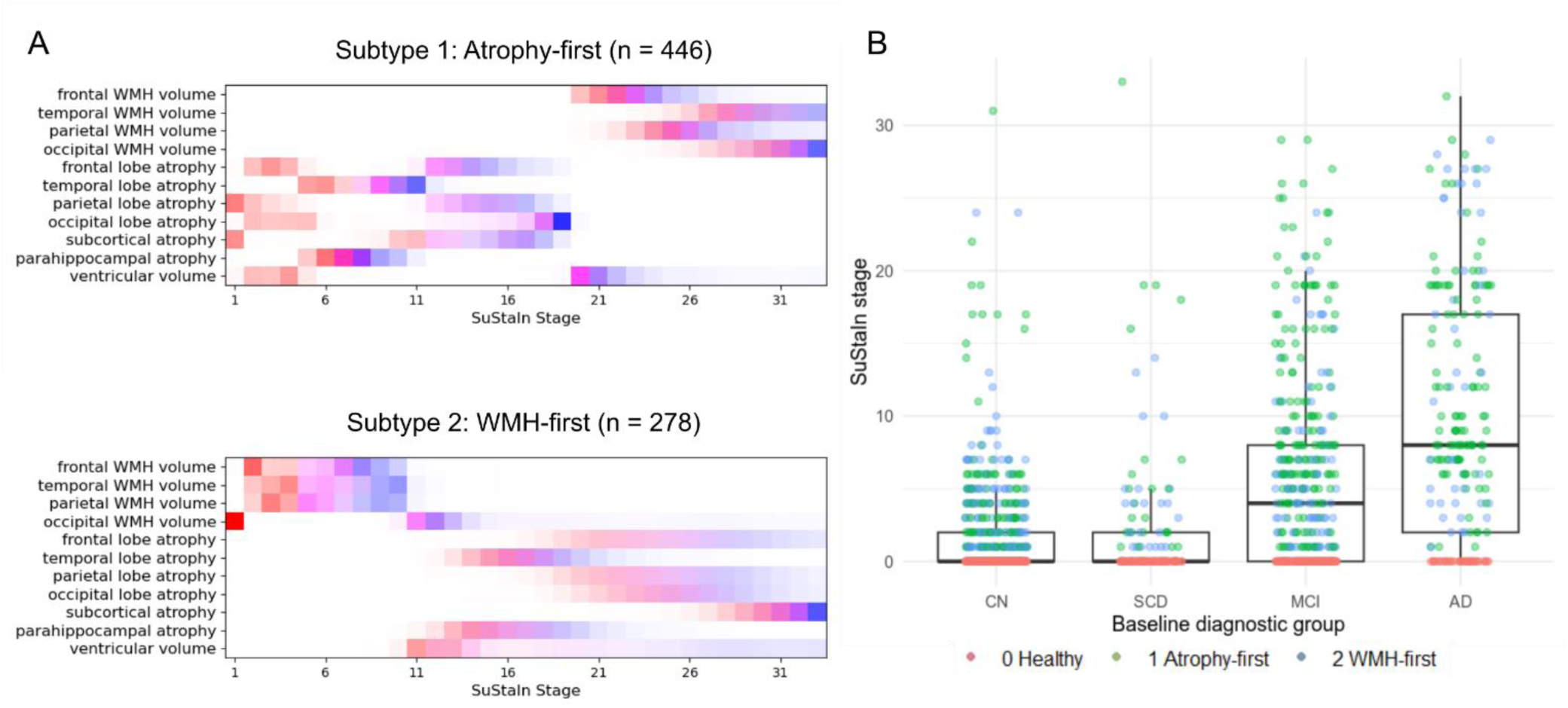
Results of the two-subtype SuStaIn model. The positional variance diagrams show the biomarker trajectories for the two-subtype model. Each box represents the certainty that a biomarker has reached a certain w-score at a given SuStaIn/pseudo-stage, with colours indicating the w-score threshold (.7, 1.4, 2.1) and higher saturation representing a higher probability. B) Stage assignment of the one-subtype model across datasets, stratified by baseline clinical diagnosis. WMH: White Matter Hyperintensity, CN: cognitively unimpaired, SCD: subjective cognitive decline, MCI: mild cognitive impairment.

The two subtypes did not significantly differ in demographic variables or baseline MoCA scores (Table 2). As expected, the atrophy-first subtype had lower GM volume, whereas the WMH-first subtype had higher WMH volumes at baseline (p < .001). Compared with subtype 0, the WMH-first subtype was significantly older (p = .001). Both abnormal subtypes had a higher proportion of APOE4 carriers and lower baseline global cognition than subtype 0 (p < .001). Most CU and SCD participants were grouped into subtype 0, whereas most AD participants were categorized as atrophy-first. However, 42 participants with a diagnosis of AD at baseline were grouped into subtype 0. These participants had, on average, higher education levels, while showing lower atrophy and WMH values, as well as lower amyloid and tau burden, than AD cases grouped within the abnormal subtypes (Supplementary Figure 4).

**Table 1.** Baseline participant characteristics in the four datasets. Continuous variables are expressed as mean (standard deviation). P-values are based on ANOVA or χ2 analysis with post-hoc analysis using pairwise t-tests for continuous variables and Fisher’s exact test for categorical variables. AD: Alzheimer’s Disease, MCI: Mild Cognitive Impairment, SCD: Subjective Cognitive Decline, CU: Cognitively Unimpaired, WMH: White Matter Hyperintensity, GM: Grey Matter, MoCA: Montreal Cognitive Assessment.

|  | ADNI | ALLFTD | CIMA-Q | PREVENT-AD | p |
| --- | --- | --- | --- | --- | --- |
| N Participants (n(%)) | 573 (41.0) | 245 (17.5) | 246 (17.6) | 333 (23.8) |  |
| Diagnosis (n(%)) |  |  |  |  | <0.001 |
| CU | 115 (20.1)<sup>c,d,e</sup> | 170 (69.4)<sup>b,d,e</sup> | 40 (16.3)<sup>b,c,e</sup> | 333 (100.0)<sup>b,c,d</sup> |  |
| SCD | 20 (3.5)<sup>c,d,e</sup> | 0<sup>b,d,e</sup> | 125 (50.8)<sup>b,c,e</sup> | _a |  |
| MCI | 266 (46.4)<sup>c,d,e</sup> | 66 (26.9)<sup>b,d,e</sup> | 64 (26.0)<sup>b,c,e</sup> | 0<sup>b,c,d</sup> |  |
| AD | 172 (30.0)<sup>c,d,e</sup> | 9 (3.7)<sup>b,d,e</sup> | 17 (6.9)<sup>b,c,e</sup> | 0<sup>b,c,d</sup> |  |
| Age | 73.7 (7.4)<sup>c,e</sup> | 62.8 (6.5)<sup>b,d</sup> | 73.0 (5.4)<sup>c,e</sup> | 63.0 (5.0)<sup>b,d</sup> | <0.001 |
| Sex (n(%)) |  |  |  |  | <0.001 |
| Female | 263 (45.9)<sup>c,d,e</sup> | 143 (58.4)<sup>b,e</sup> | 154 (62.6)<sup>b</sup> | 234 (70.3)<sup>b,c</sup> |  |
| Male | 310 (54.1)<sup>c,d,e</sup> | 102 (41.6)<sup>b,e</sup> | 92 (37.4)<sup>b</sup> | 99 (29.7)<sup>b,c</sup> |  |
| Education (years) | 15.7 (2.8) | 15.8 (2.7) | 15.1 (3.5) | 15.5 (3.4) | 0.052 |
| APOE ε4 alleles (n(%)) | | | | | <0.001 |
| 0 | 252 (44.7)<sup>d,e</sup> | _a | 140 (92.7)<sup>b,e</sup> | 202 (61.0)<sup>b,d</sup> |  |
| 1 | 232 (41.1)<sup>d,e</sup> | _a | 3 (2.0)<sup>b,e</sup> | 121 (36.6)<sup>b,d</sup> |  |
| 2 | 80 (14.2)<sup>d,e</sup> | _a | 8 (5.3)<sup>b,e</sup> | 8 (2.4)<sup>b,d</sup> |  |
| MoCA total score | 21.6 (4.8)<sup>c,d,e</sup> | 26.1 (5.5)<sup>b,e</sup> | 25.9 (3.5)<sup>b,e</sup> | 28.0 (1.6)<sup>b,c,d</sup> | <0.001 |
| Total WMH volume (mm<sup>3</sup>) | 5892.6 (8234.7)<sup>c,d,e</sup> | 2775.3 (2871.4)<sup>b,d,e</sup> | 4485.6 (5853.1)<sup>b,c,e</sup> | 2322.1 (3167.5)<sup>b,c,d</sup> | <0.001 |
| Total GM volume (mm<sup>3</sup>) | 416080.2 (59982.7)<sup>c,d,e</sup> | 456067.7 (70425.0)<sup>b,e</sup> | 463571.2 (62282.8)<sup>b</sup> | 471353.2 (65167.4)<sup>b,c</sup> | <0.001 |

<sup>a</sup> Information not available for the majority of participants at baseline.
<sup>b</sup> Significantly different from ADNI.
<sup>c</sup> Significantly different from ALLFTD.
<sup>d</sup> Significantly different from CIMA-Q.
<sup>e</sup> Significantly different from PREVENT-AD.

**Table 2.** Baseline characteristics of participants in distinct SuStaIn subtypes in the four datasets. Continuous variables are expressed as mean (standard deviation). P-values are based on ANOVA or χ2 analysis. AD: Alzheimer’s Disease, MCI: Mild Cognitive Impairment, SCD: Subjective Cognitive Decline, CU: Cognitively Unimpaired, WMH: White Matter Hyperintensity, MoCA: Montreal Cognitive Assessment, CSF: cerebrospinal fluid.

|  | Healthy subtype (subtype 0) | Atrophy-first | WMH-first | p |
| --- | --- | --- | --- | --- |
| N Participants (n(%)) | 671 (48.0) | 448 (32.1) | 278 (19.9) |  |
| Diagnosis (n(%)) |  |  |  | <0.001 |
| CU | 355 (52.9)<sup>b,c</sup> | 127 (28.3)<sup>a,c</sup> | 102 (36.7)<sup>a,b</sup> |  |
| SCD | 97 (14.5)<sup>b,c</sup> | 24 (5.4)<sup>a,c</sup> | 24 (8.6)<sup>a,b</sup> |  |
| MCI | 177 (26.4)<sup>b,c</sup> | 187 (41.7)<sup>a,c</sup> | 106 (38.1)<sup>a,b</sup> |  |
| AD | 42 (6.3)<sup>b,c</sup> | 110 (24.6)<sup>a,c</sup> | 46 (16.5)<sup>a,b</sup> |  |
| Age | 68.4 (8.1)<sup>c</sup> | 69.4 (8.4) | 70.5 (8.0)<sup>a</sup> | 0.001 |
| Sex (n(%)) |  |  |  | 0.714 |
| Female | 384 (57.2) | 258 (57.6) | 152 (54.7) |  |
| Male | 287 (42.8) | 190 (42.4) | 126 (45.3) |  |
| Education (years) | 15.4 (3.1) | 15.7 (2.9) | 15.9 (3.2) | 0.101 |
| MoCA total score | 26.7 (3.1)<sup>b,c</sup> | 24.3 (6.6)<sup>a</sup> | 24.9 (4.5)<sup>a</sup> | <0.001 |
| APOE ε4 alleles (n(%)) | | | | <0.001 |
| 0 | 325 (67.3)<sup>b,c</sup> | 167 (46.0)<sup>a</sup> | 102 (51.0)<sup>a</sup> |  |
| 1 | 139 (28.8)<sup>b,c</sup> | 146 (40.2)<sup>a</sup> | 71 (35.5)<sup>a</sup> |  |
| 2 | 19 (3.9)<sup>b,c</sup> | 50 (13.8)<sup>a</sup> | 27 (13.5)<sup>a</sup> |  |
| Total WMH volume (mm<sup>3</sup>) | 2034.1 (1338.9)<sup>b,c</sup> | 3702.3 (5143.0)<sup>a,c</sup> | 10469.8 (10166.3)<sup>a,b</sup> | <0.001 |
| Total Grey Matter Volume (mm<sup>3</sup>) | 452966.0 (64154.6)<sup>b,c</sup> | 404260.9 (55038.9)<sup>a,c</sup> | 488998.3 (60497.9)<sup>a,b</sup> | <0.001 |

<sup>a</sup> Significantly different from Subtype 0.
<sup>b</sup> Significantly different from Subtype 1.
<sup>c</sup> Significantly different from Subtype 2.

SuStaIn stage assignments across all cohorts and subtypes were coherent with clinical diagnoses at baseline (Figure 1B), with CU and SCD participants clustering at lower stages, MCI at intermediate stages, and AD cases at higher stages. This preserved clinical gradient within biologically distinct subtypes indicates that the identified subtypes reflect qualitatively different pathological trajectories rather than differences in overall disease severity.

### 3.3. Molecular marker levels differ between subtypes

To further characterize the subtypes, we first compared cross-sectional levels of molecular markers between the SuStaIn subtypes (Figure 2). Since the available molecular measures, assay kits, and markers differed across datasets, molecular markers were not directly comparable across cohorts and could not be reliably harmonized for a pooled analysis. We therefore performed the analyses separately within each dataset. However, this approach substantially reduced the sample size available for each analysis and, consequently, statistical power to detect differences between SuStaIn subtypes. In ADNI, both the atrophy-first (β = −.55, p.adj < .001) and the WMH-first subtype (β = −.65, p.adj < .001), showed lower CSF amyloid-β42 levels than subtype 0, indicating a higher amyloid burden in the abnormal subtypes. The WMH-first subtype also had slightly higher SUVr values in amyloid PET than subtype 0 (β = −.38, p = .04, p.adj = .12), although this effect did not survive FDR correction. Furthermore, CSF p-tau levels were increased in both abnormal subtypes (atrophy-first: β = .33, p = .02; WMH-first: β = .33, p = .04) than the healthy subtype. CSF total tau was only increased in subtype 1 (β = .33, p = .02). In CIMA-Q, comparisons were only significant before FDR correction. The atrophy-first subtype showed lower CSF amyloid-β42 (β = −.62, p = .05, p.adj = .14) and amyloid-β40 (β = −.81, p = .05) than subtype 0. The WMH-first subtype exhibited higher plasma p-tau231 values than subtype 0 ( β = .41, p = .04, p.adj = .11).

**Figure 2.**
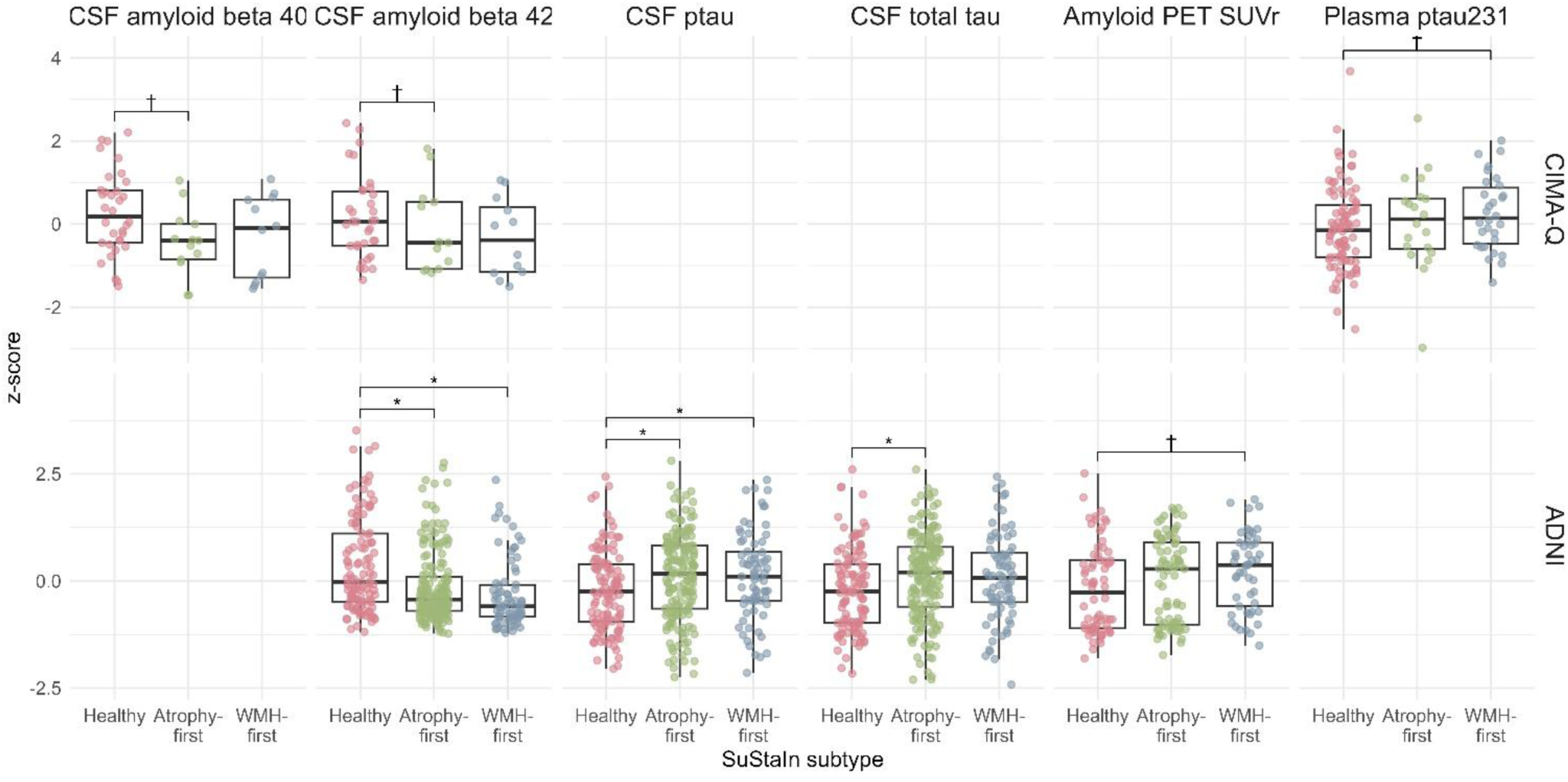
Baseline levels of molecular markers across SuStaIn subtypes, stratified by dataset. Boxplots show levels of CSF/plasma/PET SUVr markers for amyloid beta and total/p-tau. Values are shown as z-scores. Asterisks indicate significant differences between subtypes based on linear regression models adjusting for age and sex after FDR-correction, while † denotes significance before FDR correction.

### 3.4. Vascular risk factors

The SuStaIn subtypes also differed in vascular risk factors at baseline (Figure 3). The atrophy-first subtype showed lower BMI (β = ‡0.17, p.adj = .03) and higher HDL/LDL cholesterol ratio (β = 0.76, p.adj < .001) compared to subtype 0. The WMH-first subtype showed elevated diastolic (β = 0.33, p.adj = .001) blood pressure relative to subtype 0 and the atrophy-first subtype (β = 0.28, p.adj = .01), as well as higher systolic blood pressure compared to the healthy subtype (β = 0.18, p.adj = .05) and the atrophy-first subtype (β = 0.22, p = .04, p.adj = .05). The HDL/LDL cholesterol ratio was also elevated in the WMH-first subtype (β = 0.37, p.adj = .01) compared to subtype 0. Additionally, the WMH-first subtype reported more cigarettes smoked per day (past or current) than the atrophy-first subtype (β = 0.35, p = .03, p.adj = .09), although this effect did not survive FDR correction. Total cholesterol, and years of smoking did not significantly differ between subtypes. Similarly, the proportion of participants with diabetes or with a history of current or past smoking did not differ among subtypes.

**Figure 3.**
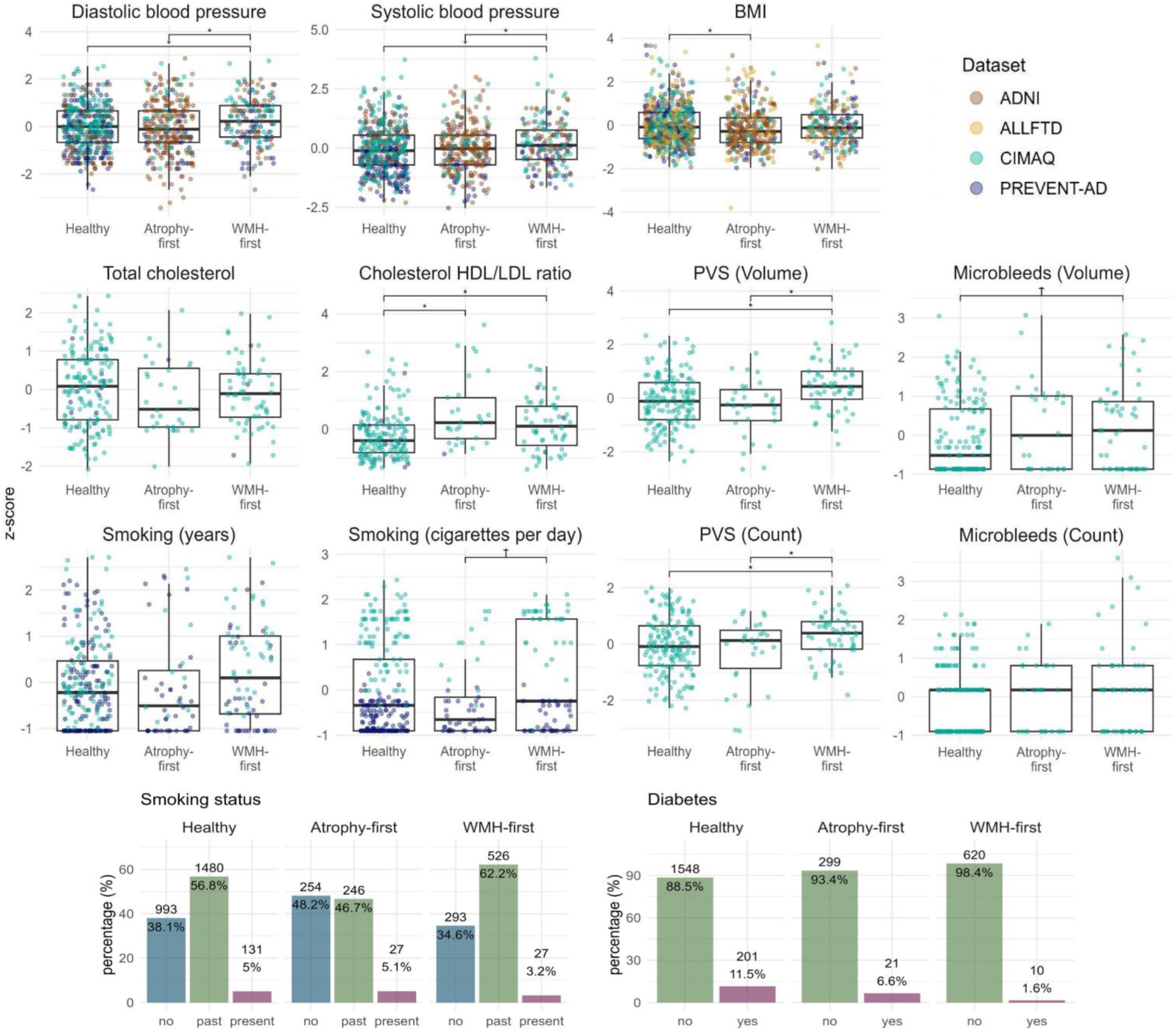
Vascular risk factors at baseline visit in each SuStaIn subtype. Values are z-scored. Colours denote the dataset of each participant. Group differences were assessed using linear regression, adjusted for age and sex and corrected for multiple comparisons using FDR correction. Asterisks (*) indicate significant differences after FDR correction, while † denotes significance before FDR correction. Categorical variables (smoking status and diabetes) are shown in bar plots. BMI: Body Mass Index, PVS: Perivascular Spaces, HDL: high-density lipoprotein, LDL: low-density lipoprotein.

PVS and microbleed segmentations were available at baseline for 241 participants in CIMA-Q (healthy subtype n = 158, atrophy-first subtype n = 30, WMH-first subtype n = 53). After adjusting for age and sex, the WMH-first subtype showed increased PVS volumes compared to both the healthy subtype (β = 0.48, p.adj = .01) and the atrophy-first subtype (β = 0.72, p.adj = .01). PVS count and microbleed count and volume did not significantly differ between groups.

### 3.5. Subtype differences in longitudinal cognitive trajectories

Longitudinal cognitive data were available for 1141 participants (healthy subtype n = 543, atrophy-first subtype n = 366, WMH-first subtype n = 232), with up to eight visits per participant (mean = 3.2 visits). The mean follow-up duration was 1.1 years, with a maximum follow-up of 3.5 years.

To assess the potential differences in trajectories of cognitive impairment across the data-driven subtypes, we compared rates of decline in cognitive tests in each dataset using linear mixed-effects models, whereby the interaction term *SuStaIn subtype x Time from baseline* denotes the difference in the slope of cognitive performance between the subtypes contrasted against the ‘healthy’ subtype 0 (Figure 4).

**Figure 4.**
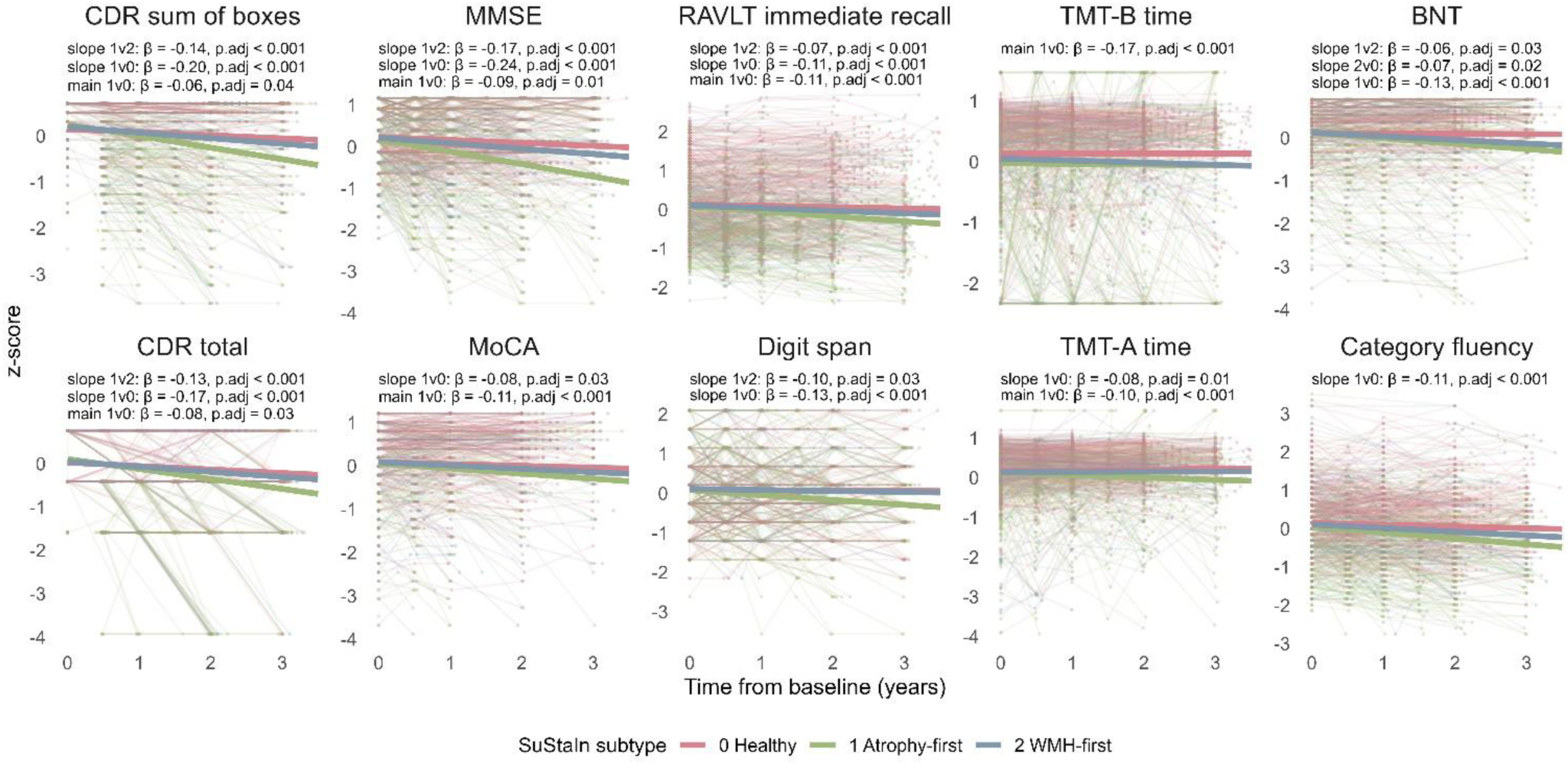
Longitudinal cognitive trajectories across SuStaIn subtypes in each dataset. Models were adjusted for age at baseline, sex, education, and baseline cognitive score. Values expressed as z-scores and adjusted consistently so that lower values correspond to worse performance. RBANS: Repeatable Battery for the Assessment of Neuropsychological Status, RAVLT: Rey Auditory Verbal Learning Test, TMT: Trail Making Test.

Compared with the healthy subtype, the atrophy-first subtype showed significantly impaired baseline cognitive performance across measures of global cognition, executive function, and memory, including MoCA (β = ‡0.10, p.adj = .01), MMSE (β = ‡0.12, p.adj < .001), TMT Part B (β = ‡0.12, p.adj = .02), and RAVLT immediate recall (β = ‡0.09, p.adj < .001). The atrophy-first subtype also demonstrated steeper cognitive decline across several domains, including global cognition measured by MoCA (β = ‡0.08, p.adj = .04), MMSE (β = ‡0.24, p.adj < .001), CDR sum of boxes (β = ‡0.20, p.adj < .001), and CDR total score (β = ‡0.17, p.adj < .001); attention/processing speed assessed through TMT Part A (β = ‡0.08, p.adj < .001) and digit span (β = ‡0.12, p.adj < .001); memory measured by RAVLT immediate recall (β = ‡0.10, p.adj < .001); and language indexed by BNT (β = ‡0.13, p.adj < .001) and category fluency (β = ‡0.11, p.adj < .001).

The WMH-first subtype showed significantly worse baseline performance on MMSE (β = ‡0.09, p.adj = .03) than subtype 0, indicating worse global cognition. This group also exhibited faster decline in language, as measured through the BNT (β = ‡0.07, p.adj = .02).

We also found significant differences between the abnormal subtypes. The atrophy-first subtype demonstrated significantly faster decline in global cognition, memory, attention, and language, as indicated by steeper slopes on the MMSE (β = ‡0.17, p.adj < .001), CDR sum of boxes (β = ‡0.14, p.adj < .001), CDR total score (β = ‡0.13, p.adj < .001), RAVLT immediate recall (β = ‡0.07, p.adj < .001), digit span (β = ‡0.09, p.adj = .03), and BNT (β = ‡0.06, p.adj = .03).

### 3.6. Subtype difference in clinical conversion rates

Multinomial logistic regression was used to examine the association between subtype and conversion group, adjusting for age and sex. 1145 participants (healthy subtype n = 553, atrophy-first subtype n = 360, WMH-first subtype n = 232) with at least one clinical follow-up to confirm diagnosis were included in this analysis (Figure 5). Relative to the healthy group, the atrophy-first subtype was associated with increased odds of stable MCI (OR = 4.04, 95% CI [2.57-6.34], p.adj < .001), conversion to AD (OR = 11.79, 95% CI [6.94-20.04], p.adj < .001), and AD (OR = 9.73, 95% CI [5.92-16.03], p.adj < .001). Individuals in the WMH-first subtype had higher odds of stable SCD (OR = 0.34, 95% CI [0.13-0.89], p.adj = .05), stable MCI (OR = 2.16, 95% CI [1.34-3.48], p.adj < .001) and AD (OR = 2.97, 95% CI [1.72-5.13], p.adj < .001).

**Figure 5.**
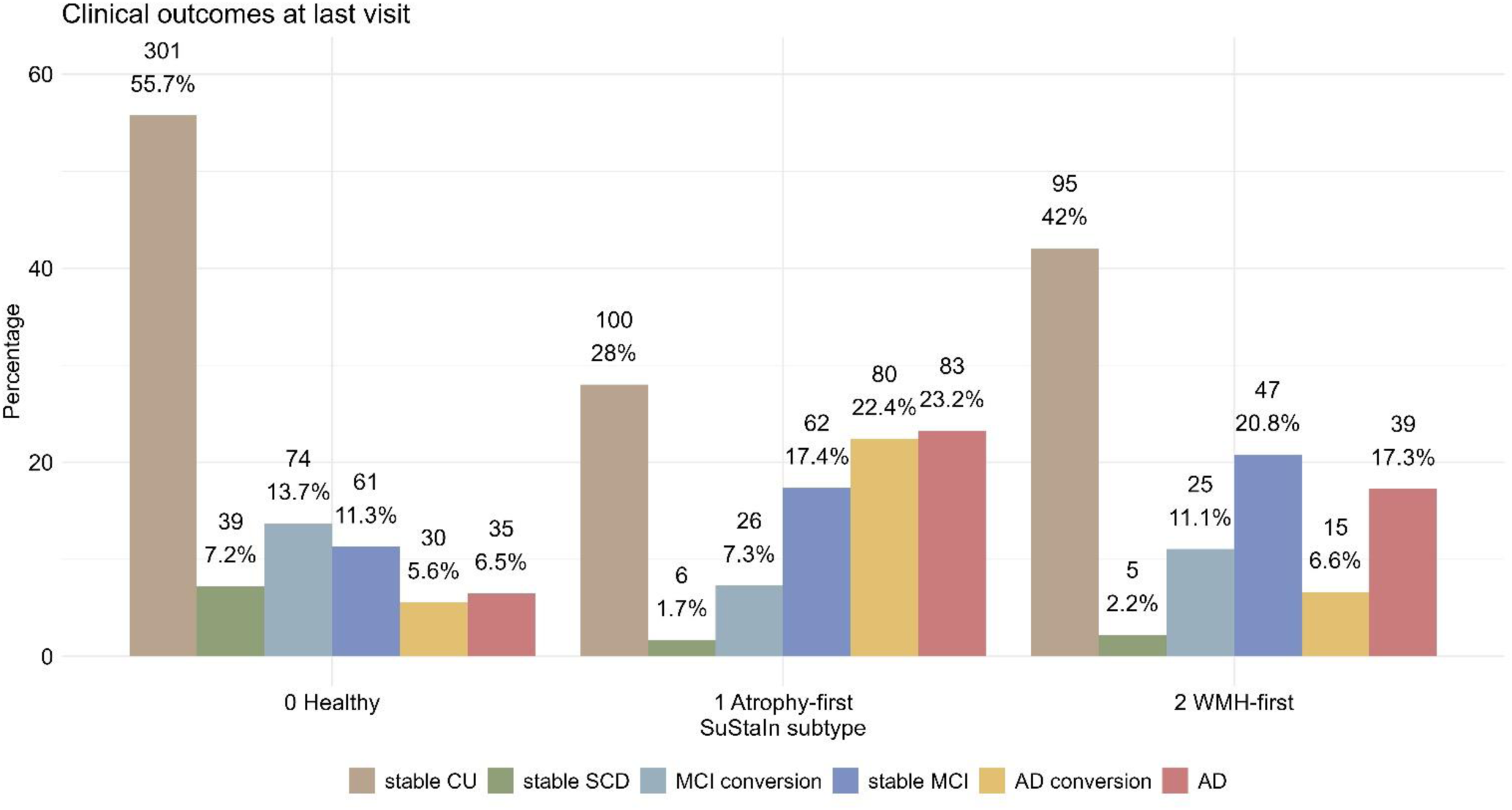
Clinical outcomes across SuStaIn subtype at the last available visit. Percentages indicate the percentage of stable cognitively unimpaired (CU), stable subjective cognitive decline (SCD), stable mild cognitive impairment (MCI), AD, conversions from CU/SCD to MCI, or from CU/SCD/MCI to AD within the respective subtype.

When WMH-first was compared with atrophy-first, the odds of AD conversion were higher for atrophy-first (OR = 0.12, 95% CI [0.06-0.23], p.adj < .001), as were the odds of AD (OR = 0.31, 95% CI [0.18-0.52], p.adj < .001). However, the WMH-first subtype was associated with higher odds of stable MCI compared with atrophy-first (OR = 0.54, 95% CI [0.32-0.89], p.adj = .03).

### 3.7. Neuropathological outcomes

Autopsy data was available for 21 participants in the atrophy-first subtype, 13 for WMH-first, and 7 of subtype 0 (Figure 6). Neuropathological outcomes differed across clinical subtypes after adjustment for age at death and sex. Compared with the healthy subtype, subtype 1 was associated with higher Braak stages (OR = 1755.57, 95% CI [33.50, 92,010.85], p < .001, p.adj < .001) and intermediate CAA (OR = 42.73, 95% CI [1.31, 1391.34], p = .03, p.adj = .35). The atrophy-first subtype also showed higher odds of TDP43 pathology than subtype 0 (OR = 10.47, 95% CI [1.06, 102.98], p = .04, p.adj = .20) and the WMH-first subtype (OR = 0.14, 95% CI [0.02, 0.82], p = .03, p.adj = .11). In contrast, the atrophy-first was associated with slightly lower odds of intermediate (OR = 0.003, 95% CI [1.02 × 10⁻⁵, 0.75], p = .04, p.adj = .13) and high arteriolosclerosis (OR = 0.003, 95% CI [1.04 × 10⁻⁵, 0.70], p = .04, p.adj = .13).

**Figure 6.**
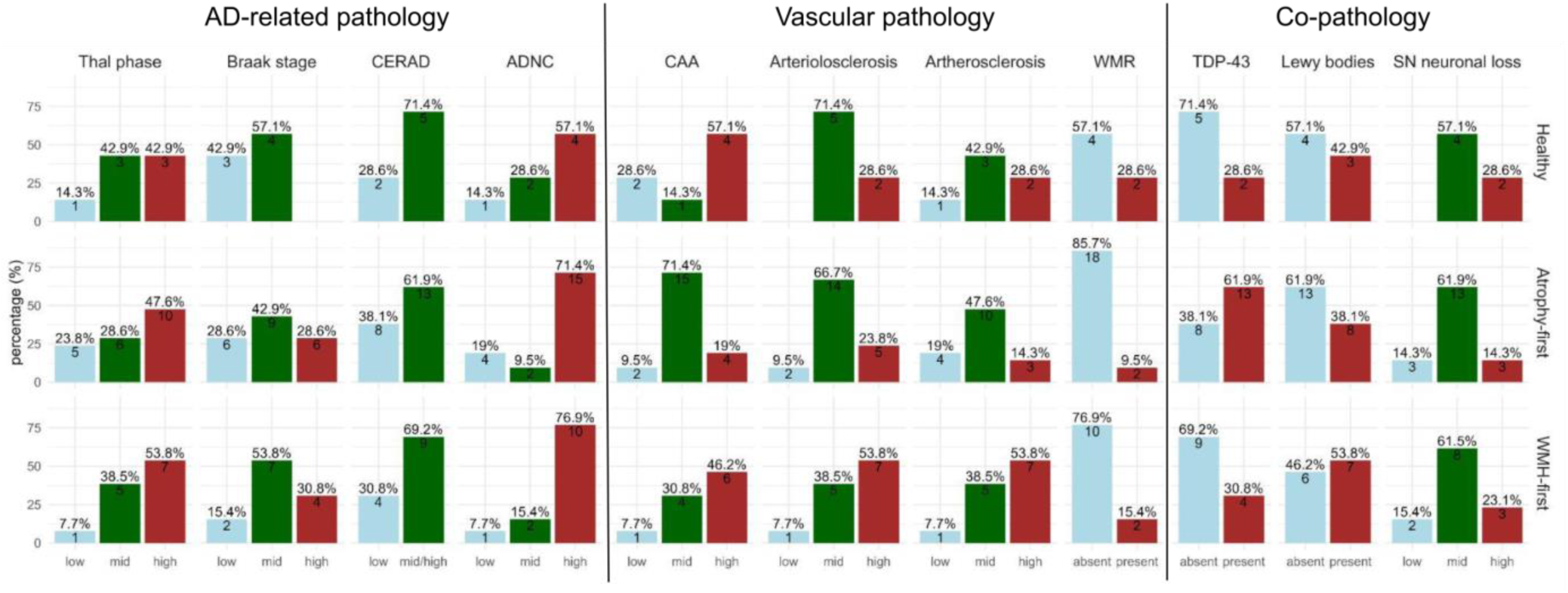
Neuropathological outcomes across SuStaIn subtypes. Autopsy data were available for 41 participants from CIMA-Q and ADNI. Pathological burden was categorized using median splits into low, intermediate, and high levels of pathology, or as present versus absent, as appropriate. Percentages indicate the proportion of participants within each SuStaIn subtype falling into the respective pathology category. Asterisks indicate significantly different odds ratios (ORs) for a given pathology level relative to the low/absent reference level between two SuStaIn subtypes, based on logistic regression, adjusting for age at death and sex. CERAD: CERAD neuritic placque score, ADNC: Alzheimer’s Disease Neuropathologic Change score, CAA: Cerebral Amyloid Angiopathy, atherosclerosis: atherosclerosis of the circle of Willis, WMR: White matter rarefaction, SN: substantia nigra.

The WMH-first subtype was similarly associated with higher odds of high Braak stage (OR = 5566.20, 95% CI [50.59, 612,380.00], p < .001, FDR < .001). Furthermore, it was also associated with higher odds of intermediate arteriolosclerosis (OR = 0.001, 95% CI [1.90 × 10⁻⁶, 0.61], p = .03, p.adj = .13), and a tendency for higher arteriolosclerosis, which did not reach statistical significance (OR = 0.004, 95% CI [6.96 × 10⁻⁶, 2.14], p = .08, p.adj = .21).

## 4. Discussion

In this study, we performed subtyping and staging analyses in four separate datasets (ALLFTD, PREVENT-AD, CIMA-Q, and ADNI) encompassing participants across the AD spectrum, including at-risk populations, individuals with MCI, and AD patients. Combining independent cohorts substantially improves diversity of representation and provides a more robust empirical basis for analyses. We stratified participants based on trajectories of regional MRI measures indexing neurodegeneration and WMH burden and identified two distinct subtypes: one initially characterized by brain atrophy, and a second presenting first with increased WMH volumes. Both subtypes were differentiated from the ‘healthy’ group by abnormal CSF or PET amyloid-β and/or tau levels at baseline. This was confirmed in the subsample with post-mortem data, where both abnormal subtypes exhibited higher Braak stages than the healthy subtype. At baseline, the WMH-first subtype was furthermore characterized by higher vascular risk burden than the healthy subtype. The atrophy- and WMH-first subtypes showed worse longitudinal cognitive decline across multiple domains compared to the healthy subtype. Additionally, the atrophy-first subtype exhibited worse longitudinal cognitive performance than the WMH-first subtype. Finally, subtypes differed in clinical outcomes and subsequent diagnoses, with the atrophy-first subtype showing a higher risk of AD or conversion to AD and the WMH-first having higher odds for MCI.

The results of this study reinforce the view that AD is not a single, homogeneous disorder but rather reflects distinct biological processes involving neurodegeneration, proteinopathy, and vascular injury, emerging along distinct trajectories^3,14,38^. The finding of an atrophy-first and a WMH-first subtype aligns with models proposing parallel but interacting pathways to cognitive decline^4,5,39^. The atrophy-dominant subtype reflects a neurodegenerative cascade, whereas the WMH-dominant subtype points to vascular contributions that may independently accelerate cognitive impairment while interacting with AD pathology.

While WMH can arise from multiple pathophysiological processes, the literature suggests that WMHs in AD result from a combination and vascular and AD-related pathologies.^40^ The vascular contribution to WMH in this sample is supported by the higher levels of blood pressure and smoking exposure observed in the WMH-first subtype compared with both other subtypes, as both are established risk factors for WMH burden.^41,42^ Furthermore, the WMH-first subtype also showed increased burden of other cerebrovascular lesions (namely dilated perivascular spaces), indicative of a vascular disease phenotype. Consistent with this interpretation, among participants with post-mortem data, the WMH-first subtype showed greater arteriolosclerosis compared with the atrophy-first subtype. In contrast, the atrophy-first subtype exhibited higher levels of CAA and TDP-43 pathology, suggesting that co-pathology may contribute to the observed atrophy pattern. Notably, both imaging-derived subtypes exhibited abnormal CSF amyloid-β and tau levels as well as post-mortem tau pathology burden, suggesting that neurodegenerative and vascular pathways may involve core AD proteinopathy even when their initial manifestations differ. This is consistent with biomarker staging models in which amyloid and tau abnormalities precede and potentially amplify both cortical atrophy and white matter injury, pointing to a shared disease process^3,40,43^.

Analysis of longitudinal cognitive trajectories across the data-driven subtypes revealed that the atrophy-first subtype was associated with steeper decline across multiple cognitive domains, including memory, executive function, attention, and global cognition, consistent with the expected pattern of AD-related neurodegeneration relative to healthy aging. This partially contrasts the existing literature, which has shown a strong contribution of WMHs specifically to executive dysfunction and processing speed^44^. Nonetheless, our results provide evidence that vascular burden contributes to cognitive decline as the WMH-first subtype largely performed worse relative to the healthy subtype across global cognition and language. This aligns with earlier findings linking WMH burden to impaired language performance specifically in MCI,^45^ which also was the most prevalent clinical diagnosis of participants grouped into our WMH-first subtype. From a clinical standpoint, the finding that the conversion rates to MCI and AD differed between the WMH-first subtype and the atrophy-first groups has important implications for individualized risk prediction, particularly given that these subtypes are already detectable in the pre-clinical stage.

While our findings align with a growing body of work applying data-driven disease progression modelling to AD^14^, direct comparisons across studies are difficult as clustering frameworks, model parameters, and input features vary widely across the literature. Several prior SuStaIn-based studies, for instance, focused exclusively on MRI-derived atrophy^15,46^ or tau PET^16^, relied on global rather than regional measures^39^, and rarely incorporated markers of vascular burden. In comparison to Mastenbroek et al. (2024), who incorporated atrophy, WMH, and fluid markers, our findings similarly reveal a distinction between a more AD-typical atrophy-driven progression and a vascular or mixed entity. The finding of two stable subtypes across ALLFTD, PREVENT-AD, CIMA-Q, and ADNI further strengthens the argument that these represent true biological phenotypes rather than dataset-specific effects.

While the cohorts analysed in this study aim to acquire data for AD or at-risk individuals, they differ in their recruitment strategies and clinical contexts. PREVENT-AD specifically recruited individuals with familial AD risk due to a first-degree family history of AD, while CIMA-Q enriched its cohort for individuals with SCD, which is known to elevate the risk of conversion to AD. ADNI comparatively includes more APOE ε4 carriers. However, ADNI’s exclusion of participants with Hachinski scores above 4 is particularly relevant to the present study, as it likely results in an underrepresentation of cerebrovascular pathology that is not representative of ageing populations. Given PREVENT-AD’s explicit focus on the pre-clinical stage of AD, PREVENT-AD participants were notably younger and showed, on average, lower disease severity than those in CIMA-Q and ADNI. Additional differences in geographic and healthcare context should be noted, as PREVENT-AD and CIMA-Q recruited exclusively from Quebec, while ALLFTD and ADNI drew from a broader North American sample with potentially different healthcare access and population characteristics. Quebec’s publicly funded universal healthcare system provides relatively equitable access to medical care, while the US operates within a more heterogeneous healthcare landscape, which could introduce systematic differences in the clinical profiles of participants.

A key strength of the present study is the consistent application of the same MRI processing pipeline, PELICAN, to derive atrophy measures and WMH volumes across datasets. PELICAN was developed specifically to handle multi-centre data and has been widely applied to a range of cohorts and populations^41,47–51^. Using the same pipeline across all datasets avoids bias or inconsistencies in this initial step. The inclusion of four independent longitudinal cohorts, together including a large sample of 1396 participants, with up to 3.5 years of follow-up duration further strengthens confidence in the observed results.

However, several limitations need to be considered. Plasma and CSF biomarker kits differed across cohorts, which may introduce variability in biomarker estimates, and ALLFTD did not collect fluid biomarkers for amyloid and tau. Consequently, we examined molecular biomarkers separately for each cohort, leading to decreased statistical power. This limitation, along with the incomplete availability of molecular biomarkers also prevented us from including these biomarkers in our subtyping model. Furthermore, cognitive follow-up assessments were not administered consistently across all visits in PREVENT-AD, and CIMA-Q did not collect cognitive follow-up data for its healthy control and AD subgroups, restricting longitudinal cognitive analyses in those samples. Despite the increased generalizability afforded by accumulating data from four separate, diverse cohorts, our sample still predominantly included White individuals with above-average education levels, which is not representative of the broader population, particularly within these age ranges.

Overall, the present study demonstrates that biologically distinct subtypes of preclinical AD can be reliably identified from multimodal imaging data and replicated across independent cohorts. The consistent emergence of atrophy-first and WMH-first trajectories underscores the importance of accounting for both neurodegenerative and vascular contributions when characterizing disease risk, and highlights the potential of data-driven subtyping approaches to refine our understanding of the underlying biological mechanisms within the AD spectrum.

## Data and Code Availability

ADNI and ALLFTD data are available under https://ida.loni.usc.edu/. CIMA-Q data can be accessed at http://www.cima-q.ca/en/home/. PREVENT-AD data can be accessed under https://openpreventad.loris.ca/. PELICAN is available at https://github.com/VANDAlab/Preprocessing_Pipeline. Codes used for analysis are available on GitHub (https://github.com/ameliemetz/AD_subtyping).

## Data Availability

Data used in preparation of this article were obtained from the Alzheimer's Disease Neuroimaging Initiative (ADNI) database (https://adni.loni.usc.edu). As such, the investigators within the ADNI contributed to the design and implementation of ADNI and/or provided data but did not participate in the analysis or writing of this report. Data used in preparation of this article were obtained from the PResymptomatic EValuation of Experimental or Novel Treatments for Alzheimer's Disease (PREVENTAD) program, data release 8.1 (https://www.centrestopad.com/) Data used in the preparation of this article were obtained from the Consortium for the early identification of Alzheimer's disease Quebec (CIMA-Q; https://cima-q.ca). A list of researchers involved in the design of CIMA-Q can be found on the cima-q.ca website. These researchers contributed to the establishment of protocols, the implementation of the research infrastructure, the recruitment and follow-up of participants, the obtaining of data, the maintenance of biological and ex-vivo samples, and certain derived data.

https://www.allftd.org/data

## Acknowledgements

The authors acknowledge Digital Research Alliance of Canada (https://www.alliancecan.ca/en) for the usage of the computing resources in the current work.

Data collection and sharing for this project was funded by the Alzheimer’s Disease Neuroimaging Initiative (ADNI) (National Institutes of Health Grant U01 AG024904) and DOD ADNI (Department of Defense award number W81XWH-12-2-0012). ADNI is funded by the National Institute on Aging, the National Institute of Biomedical Imaging and Bioengineering, and through generous contributions from the following: AbbVie, Alzheimer’s Association; Alzheimer’s Drug Discovery Foundation; Araclon Biotech; BioClinica, Inc.; Biogen; Bristol-Myers Squibb Company; CereSpir, Inc.; Cogstate; Eisai Inc.; Elan Pharmaceuticals, Inc.; Eli Lilly and Company; EuroImmun; F. Hoffmann-La Roche Ltd and its affiliated company Genentech, Inc.; Fujirebio; GE Healthcare; IXICO Ltd.; Janssen Alzheimer Immunotherapy Research & Development, LLC.; Johnson & Johnson Pharmaceutical Research & Development LLC.; Lumosity; Lundbeck; Merck & Co., Inc.; Meso Scale Diagnostics, LLC.; NeuroRx Research; Neurotrack Technologies; Novartis Pharmaceuticals Corporation; Pfizer Inc.; Piramal Imaging; Servier; Takeda Pharmaceutical Company; and Transition Therapeutics. The Canadian Institutes of Health Research is providing funds to support ADNI clinical sites in Canada. Private sector contributions are facilitated by the Foundation for the National Institutes of Health (www.fnih.org). The grantee organization is the Northern California Institute for Research and Education, and the study is coordinated by the Alzheimer’s Therapeutic Research Institute at the University of Southern California. ADNI data are disseminated by the Laboratory for Neuro Imaging at the University of Southern California.

Data collection and dissemination of the data presented in this manuscript was supported by the ALLFTD Consortium (U19: AG063911, funded by the National Institute on Aging and the National Institute of Neurological Diseases and Stroke) and the former ARTFL & LEFFTDS Consortia (ARTFL: U54 NS092089, funded by the National Institute of Neurological Diseases and Stroke and National Center for Advancing Translational Sciences; LEFFTDS: U01 AG045390, funded by the National Institute on Aging and the National Institute of Neurological Diseases and Stroke). The authors acknowledge the invaluable contributions of the study participants and families and the assistance of the support staff at each of the participating sites.

The CIMA-Q investigators contributed to the design, protocols and implementation of the study, as well as the collection of clinical, cognitive and neuroimaging data and biological samples. A complete list of the CIMA-Q investigators can be found at www.cima-q.ca. The CIMA-Q is supported by the Fonds de recherche du Québec-Santé (FRQS) - Pfizer Innovation Program, FRQ cohort funds, the Quebec Network for Research on aging, the Fondation Courtois (NeuroMod project), the Consortium for the Neurodegeneration associated with Aging, CIHR and the Fondation Famille Lemaire.

Data used in the preparation of this article were obtained from the PRe-symptomatic EValuation of Experimental or Novel Treatments for Alzheimer’s Disease (PREVENT-AD) program data release 8.0. PREVENT-AD was launched in 2011 as a $13.5 million, 7-year public-private partnership using funds provided by McGill University, the Fonds de Recherche du Québec – Santé (FRQ-356162), an unrestricted research grant from Pfizer Canada, the J.L. Levesque Foundation, the Douglas Hospital Research Centre and Foundation, the Government of Canada, the Canada Fund for Innovation, the Canadian Institutes of Health Research (SV: 178385, JP: 153287, 178210, LC: 165921, TS: 175328,) the Alzheimer Society of Canada, the National Institutes of Health of the United States (NS: NIH AG068563), the Alzheimer Association (SB: AARG-NTF 926696) and Brain Canada Foundation.

